# The Relationship Between Social Vulnerability and Obstructive Sleep Apnea Severity at Referral to a Tertiary Sleep Centre: A Retrospective Observational Study

**DOI:** 10.64898/2026.03.12.26348278

**Authors:** Nicole M. Duff, Willis H. Tsai, Emma E. M. Spence, Ada Ip-Buting, Kerry McBrien, Maoliosa Donald, Oliver David, Gabriel E. Fabreau, Marcus Povitz, Reg Gerlitz, Jaana Woiceshyn, Sachin R. Pendharkar

**Author notes:** **Corresponding Author:** Sachin R. Pendharkar, Cal Wenzel Precision Health Building, Rm 3E23, 3280 Hospital Drive NW, Calgary, Alberta, Canada, T2N 4Z6. **Authorship declaration:** Study conception: SRP, ND, ES Data acquisition: AIP Data analysis: WHT Manuscript preparation: ND, SP Critical revision and approval of the final manuscript: all authors.

## Abstract

**Rationale:** Obstructive sleep apnea (OSA) is a common, treatable chronic disease with significant health and societal consequences. Many patients face barriers to care due to systemic inequality, poverty, and other contributors to social vulnerability, leading to delayed diagnosis and more severe disease at presentation. Several studies have examined the impacts of social vulnerability on OSA severity using individual-level factors. However, there is comparatively limited work examining how neighbourhood-level indicators may influence OSA severity. This study aimed to determine whether social vulnerability, measured using a neighbourhood-level multidimensional index, is associated with OSA severity at referral to a tertiary sleep centre.

**Methods:** We conducted a retrospective observational study of adult patients referred to an academic hospital in Calgary, Canada for evaluation of OSA between November 2016 and November 2019. Patient data were linked using residential postal codes to the Canadian Index of Multiple Deprivation (CIMD), a census-based tool designed to reflect dimensions of social vulnerability in Canadian populations. CIMD divides social vulnerability into four dimensions including residential instability, ethnocultural composition, economic dependency, and situational vulnerability. We employed both linear and logistic mixed-effects models to assess the impact of neighbourhood-level social vulnerability on sleep apnea severity, using postal code as the grouping variable. OSA severity was based on home sleep apnea test (HSAT) derived oxygen desaturation index (ODI). Secondary outcomes included severe OSA (ODI ≥ 30), sleepiness based on Epworth Sleepiness Scale (ESS), and severe sleepiness (ESS > 15).

**Results:** The study included 2,232 patients, 80% of whom had at least mild OSA. ODI was positively associated with situational vulnerability (*p* < 0.01) and inversely associated with ethnocultural composition (*p* < 0.01), though both associations lost significance after adjusting for BMI. ESS was independently associated with situational vulnerability (*p* < 0.01) and inversely with ethnocultural composition (*p* = 0.01), independent of BMI and ODI. Severe sleepiness was associated with situational vulnerability (*p* < 0.01) and residential instability (*p* = 0.02).

**Conclusion:** Living in a socially deprived area was associated with OSA severity at time of referral, though this relationship appeared to be mediated by BMI. Deprivation dimensions were independently associated with sleepiness, highlighting the broader impact of social-related factors on sleepiness. These findings demonstrate the complex interplay between social vulnerability and sleep disorders and suggest that composite indices like the CIMD can enhance our understanding of these relationships.

## Introduction

Obstructive sleep apnea (OSA) is a highly prevalent and treatable chronic disease, affecting nearly one billion adults globally^1^. Untreated OSA is associated with substantial morbidity including hypertension, diabetes, cardiovascular disease, stroke, and depression^2–4^. Furthermore, it contributes to poor quality of life, increased risk of motor vehicle collisions, losses in workplace productivity, and significant financial burden on the health care system^5–7^. Despite the evidence of clinical and economic benefit of OSA treatment^5,8–10^, many patients experience delays in receiving a diagnosis and as many as 80% are undiagnosed^2,3,11–14^.

Under-diagnosis of OSA may be attributable to patient, provider, and system-level factors^12,15–18^. From a patient perspective, various social determinant of health (SDOH) have been identified as important contributors to disparities in care^12,15,16^. Previous studies have reported delayed diagnoses and increased severity of OSA in lower socioeconomic status (SES) groups^15^, ethnic/racial minorities^19^, neighborhoods with poor walkability and high crowding^16,20^, rural residence^21,22^, and those who travel further for specialist care^11^. Despite extensive exploration of these individual-level factors, there is comparatively limited work examining how broader, neighbourhood-level indicators may influence OSA severity^23^. The Canadian Index of Multiple Deprivation (CIMD) is a census-based tool designed to reflect dimensions of social vulnerability in Canadian populations^24,25^. It uses dissemination areas (DA), which are small, geographic units established by Statistics Canada and typically include 400-700 individuals^26^. The CIMD utilizes 21 variables with known associations with deprivation and marginalization, categorized into four dimensions: residential instability, ethnocultural composition, economic dependency, and situational vulnerability (Table 1)^26^. Unlike individual markers of social vulnerability, CIMD reflects the vulnerability of a patient’s surrounding environment. As a geographical index of health and social well-being, the CIMD can be used to better understand social disparities by region and identify marginalized communities^26^.

**Table 1:** The four dimensions of multiple deprivation and their corresponding indicators. *Adapted from the 2021 CIMD User Guide, Prairie Region*^26^.

|  |
| --- |
| **Residential Instability** |
| Proportion of dwellings that are apartment buildings |
| Proportion of persons living alone |
| Median 2021 household income<sup>1</sup> |
| Proportion of movers within the past 5 years |
| Proportion of population receiving government transfer payments |
| **Ethnocultural Composition** |
| Proportion of the population that is foreign born |
| Proportion of the population self-identified as visible minority |
| Proportion of the population with no knowledge of either official language (linguistic isolation) |
| Proportion of population with no religious affiliations |
| Proportion of the population which are recent immigrants |
| **Economic Dependency** |
| Proportion of the population participating in the labor force (aged 15 and older)<sup>1</sup> |
| Ratio of employment to population<sup>1</sup> |
| Dependency ratio (population aged 0-14 and aged 65 and older divided by population aged 15-64) |
| **Situational Vulnerability** |
| Proportion of the population identified as Indigenous |
| Proportion of homes needing major repairs |
| Proportion of the population aged 25-64 without a high-school diploma |
| Proportion of single parent families |
| Median dollar value of dwelling<sup>1</sup> |
| Proportion of children younger than age 6 |

<sup>1</sup> This indicator was reverse-coded, meaning it was coded opposite of the measure. For example, proportion of population that is married or common-law becomes proportion of population that is single, divorced, separated or widowed.

While there is extensive literature linking individual markers of vulnerability with OSA, most patients experience multiple, overlapping identities that shape their health^27^. This highlights the need for neighbourhood-level measures that capture the broader social context in which patients live and receive care. The aim of this study was to investigate if neighborhood-level social vulnerability, measured using a validated multidimensional index, was associated with the severity of OSA at the time of referral to a tertiary sleep center, thereby highlighting possible inequities in referral patterns or access to sleep-related care.

## Methods

### Study Design

We conducted a retrospective observational cohort study using data from patients with suspected OSA who were referred to the Foothills Medical Centre (FMC) Sleep Centre in Calgary, Canada. Ethics approval for this study was provided by the University of Calgary Conjoint Health Research Ethics Board (REB21-0433).

### Study Setting

In the province of Alberta, primary care providers (PCPs) have the option of referring patients with suspected OSA to a sleep specialist or independently managing these patients, including ordering home sleep apnea testing and initiating therapy based on the results. In the city of Calgary, patients who are referred to a sleep specialist may be referred to the FMC Sleep Centre or one of several community-based sleep specialists. Publicly funded sleep diagnostic testing is available at the FMC Sleep Centre or testing can be obtained privately at community sleep laboratories or from positive airway pressure (PAP) vendors. Patients may purchase PAP therapy or mandibular advancement devices out-of-pocket or through private insurance. Government funding is also available for seniors with low income, individuals receiving social assistance, or those who require non-invasive ventilation or oxygen therapy for more severe sleep-disordered breathing.

The FMC Sleep Centre is the sole tertiary, academic sleep centre in Southern Alberta and serves a catchment population of roughly 2.5 million. This centre receives approximately 3500 referrals annually, 46% of which are for OSA^28^. Patients who are referred to the FMC Sleep Centre undergo a triage intake process that involves home sleep apnea testing (HSAT) and a sleep questionnaire, following which they are scheduled for assessment by a sleep physician.

### Study Population

We included all patients over the age of 18 years referred to the FMC Sleep Centre with valid Alberta Health Care insurance for evaluation of OSA between November 1^st^, 2016 and November 30^th^, 2019. Patients with missing data for the primary outcomes, including HSAT and/or Epworth Sleepiness Scale, were excluded. The study cohort was derived from electronic medical record data (Figure 1).

**Figure 1:**
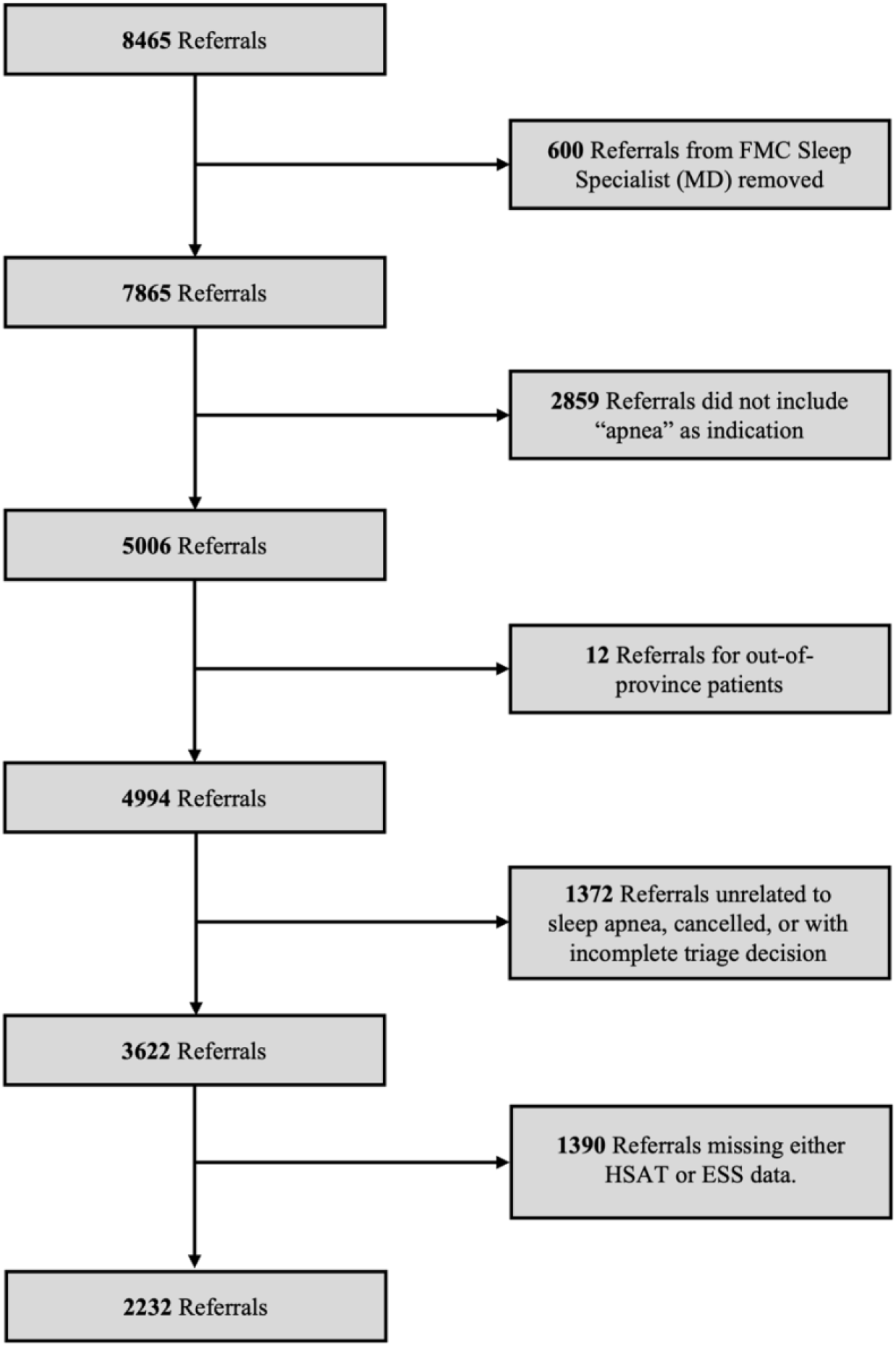
Cohort Derivation. FMC – Foothills Medical Centre, HSAT – home sleep apnea test, ESS – Epworth Sleepiness Scale

### Data Sources

During the study period, clinical documentation of OSA symptoms, medications, sleep diagnostic data, and treatment adherence records were available from the electronic medical record. These data were extracted by a data analyst at the FMC Sleep Centre and cleaned by a research associate. Comorbidities were determined using validated administrative health data algorithms^29^. Administrative data sources included the Canadian Institute for Health Information (CIHI) Discharge Abstract Database, CIHI National Ambulatory Care Reporting System, Alberta Health practitioner claims-database, and Alberta Precision Labs database.

### Exposure of Interest

The CIMD is a census-based tool designed to describe neighborhood-level social vulnerability across four dimensions: residential instability, ethnocultural composition, economic dependency, and situational vulnerability (Table 1)^24,25^. Neighborhoods are estimated by DA in the Canadian Census, similar to census block groups in the United States. Each neighborhood is assigned continuous factor scores for each dimension, which are then ranked and divided into quintiles^26^. Higher CIMD scores reflect greater disadvantage^26^. Additional census variables including age, sex, income, education, marital status, employment, and language were derived using the Postal Code Conversion File Plus (PCCF+) Version 7D (released November 2020)^30^. The PCCF+ 7D is a tool which links six-character postal codes to standard census geographic areas using a population-weighted algorithm to improve geographic assignment for residential postal codes with multiple possible census areas^30^. For this study, we used 2016 census data as most representative of the study population at the time of referral. Data linkage was performed by a geomatics analyst at the University of Calgary.

### Primary and Secondary Outcomes

The primary outcome was OSA severity, defined as oxygen desaturation index (ODI) on home sleep apnea testing (Remmers Sleep Recorder-Sagatech Electronics, Inc.) performed at the time of referral. Secondary outcomes included: the presence of severe OSA (ODI ≥ 30), daytime sleepiness based on ESS at the time of referral, and severe sleepiness (ESS > 15).

### Data Analysis

Descriptive statistics were reported on baseline characteristics including sociodemographic, comorbidities, severity of OSA as measured by ODI on home sleep apnea testing, and subjective sleepiness as measured by ESS. We employed both linear and logistic mixed-effects models to assess the association between neighbourhood-level social vulnerability and sleep apnea severity, using postal code as the grouping variable. Social vulnerability was defined using quintiles for each CIMD dimension.

Covariates included age, sex and BMI at the time of referral. Models assessing sleepiness included ODI as an additional covariate. Census data (age, sex, income, education, marital status, language, employment status) were excluded from regression models due to collinearity with CIMD data. Statistical analysis was performed using STATA version 17 (StataCorp). Results were considered statistically significant if p < 0.05.

## Results

The study cohort included 2,232 patients, for which baseline characteristics are presented in Table 2. Within our study population, 80% of patients had at least mild OSA and mean (SD) ODI was 21 (21) hr^-1^. The most prevalent comorbidities in our population included hypertension (36%), diabetes (18%), and depression (20%). Census data revealed that, compared to the Calgary and Alberta populations, the study population lived in areas with higher employment, but lower rates of post-secondary education. Median income was similar to the Calgary population but slightly higher than the Alberta population (Table 3).

**Table 2:**
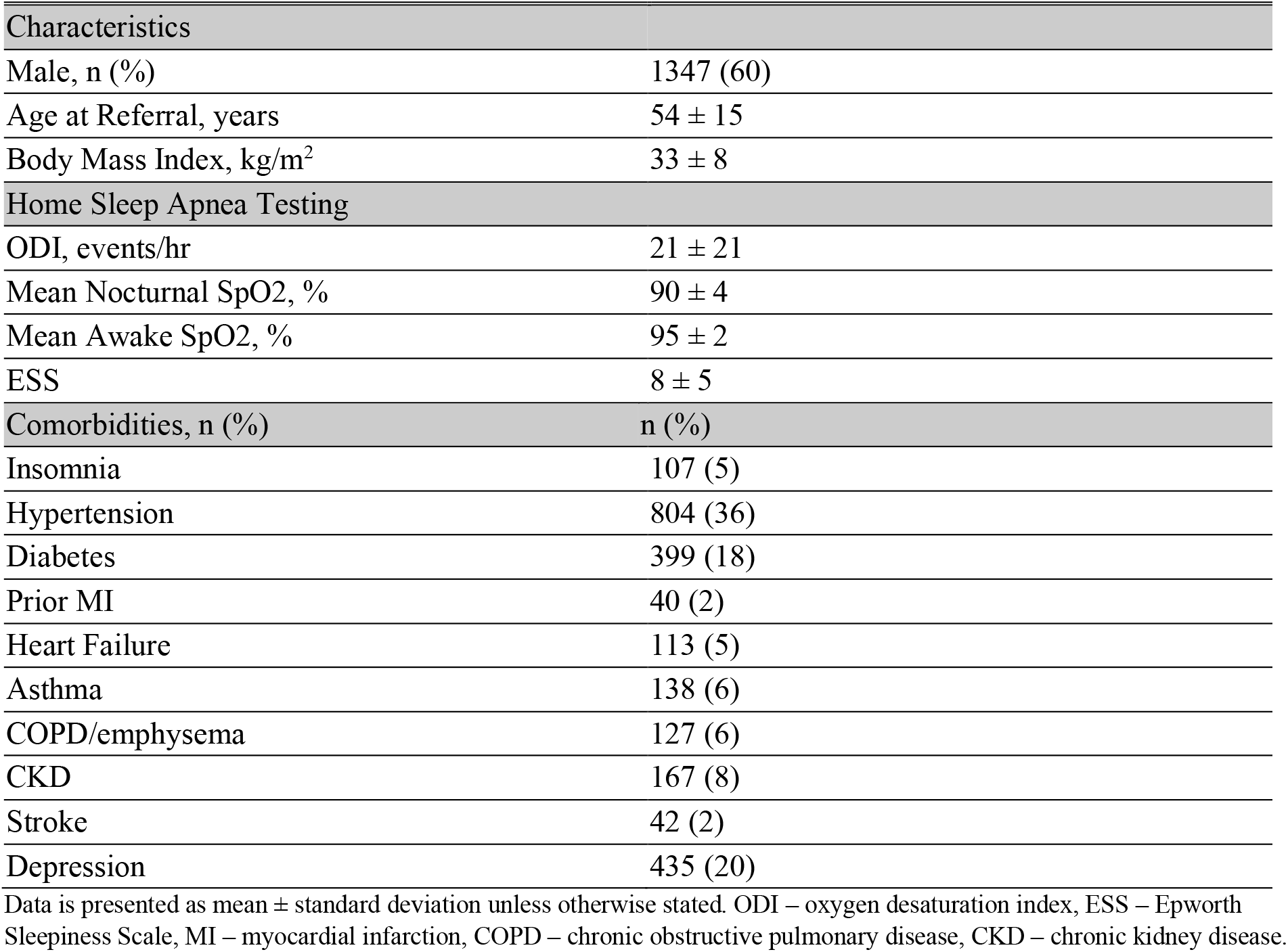
Baseline characteristics of patients included in analysis (n = 2232).

**Table 3:** 2016 Canadian Census Data for Study Population Dissemination Area compared to the City of Calgary and Province of Alberta.

| Census Variable | Study Population | City of Calgary | Province of Alberta |
| --- | --- | --- | --- |
| Male, n (%) | 49.6% | 49.9% | 50.1% |
| Median recipient income, CAD | \$43,798 | \$43,974 | \$42,717 |
| Post-secondary Education | 46.8% | 60.4% | 55.2% |
| Married or common law | 50.2% | 59.9% | 59.9% |
| Employment rate | 69.2% | 66.5% | 65.4% |

CAD – Canadian dollar

Each situational vulnerability quintile was associated with an increase in ODI of 0.99 events/hr (95% CI 0.28, 1.71, p < 0.01). In contrast, vulnerability based on ethnocultural composition was associated with a 1.19 event/hr (95% CI −2.07, −0.32) *decrease* in ODI per quintile (p < 0.01). When adjusted for BMI, these associations were no longer statistically significant. Similar results were obtained when assessing associations with severe OSA, though the inverse association between ethnocultural composition and severe OSA remained significant after adjustment (Table 4).

**Table 4:** Association between ODI and CIMD domains.

| ODI Total | Unadjusted |  | Adjusted for BMI |  |
| --- | --- | --- | --- | --- |
|  | Coefficient (95% CI) | *P value* | Coefficient (95% CI) | *P value* |
| Ethnocultural composition | - 1.19 (-2.07 - -0.32) | < 0.01 | - 0.69 (-1.53 – 0.16) | 0.11 |
| Situational vulnerability | 0.99 (0.28 – 1.71) | < 0.01 | 0.04 (-0.66 – 0.74) | 0.92 |
| Residential instability | - 0.18 (-0.80 – 0.45) | 0.58 | - 0.27 (-0.87 – 0.33) | 0.37 |
| Economic dependency | 0.54 (-0.13 – 1.21) | 0.11 | 0.56 (-0.09 – 1.20) | 0.09 |
| Severe OSA (ODI ≥ 30) | Odds ratio (95% CI) | *P value* | Odds ratio (95% CI) | *P value* |
| Ethnocultural composition | 0.88 (0.80 – 0.97) | 0.01 | 0.90 (0.81 – 0.99) | 0.03 |
| Situational vulnerability | 1.07 (0.98 – 1.15) | 0.12 | 1.00 (0.92-1.09) | 0.99 |
| Residential instability | 0.99 (0.92 – 1.06) | 0.77 | 0.97 (0.90 – 1.05) | 0.50 |
| Economic dependency | 1.05 (0.98 – 1.14) | 0.18 | 1.05 (0.97 – 1.14) | 0.22 |

CIMD – Canadian Index of Multiple Deprivation, ODI – oxygen desaturation index, OSA – obstructive sleep apnea

The analysis also revealed contrasting associations between ESS score and CIMD domains. Each situational vulnerability quintile was associated with a 0.37 point (95% CI 0.19 - 0.55, p < 0.01) increase in ESS. In contrast, each ethnocultural composition quintile was associated with a 0.27 point (95% CI - 0.48 - −0.06, p = 0.01) *decrease* in ESS. These associations were independent of BMI and ODI (Table 5). Severe sleepiness was associated with living in an area with higher situational vulnerability (p < 0.01), lower residential instability (p = 0.02), and higher ODI (p < 0.01).

**Table 5:** Sleepiness and Deprivation, with subset assessment of Severe Sleepiness (ESS > 15)

| ESS | ESS Total |  | ESS > 15 |  |
| --- | --- | --- | --- | --- |
|  | Coefficient (95% CI) | *P value* | Odds ratio (95% CI) | *P value* |
| Ethnocultural composition | -0.27 (-0.48 - -0.06) | 0.01 | 0.89 (0.79 – 1.00) | 0.05 |
| Situational vulnerability | 0.37 (0.19 – 0.55) | < 0.01 | 1.24 (1.12 – 1.37) | < 0.01 |
| Residential instability | -0.04 (-0.20 – 0.11) | 0.58 | 0.90 (0.82 – 0.98) | 0.02 |
| Economic dependence | -0.07 (-0.23 – 0.10) | 0.41 | 0.99 (0.90 – 1.10) | 0.90 |
| BMI | 0.02 (-0.01 – 0.05) | 0.16 | 1.01 (0.99 – 1.02) | 0.50 |
| ODI total | 0.04 (0.03 – 0.05) | < 0.01 | 1.02 (1.01 – 1.02) | < 0.01 |

ESS – Epworth Sleepiness Scale, BMI – body mass index, ODI – oxygen desaturation index

## Discussion

The results of this study demonstrate that in a tertiary sleep clinic referral population, there was a significant association between neighborhood-level CIMD deprivation dimensions and OSA severity. However, this association was attenuated following adjustment for BMI. We also observed an independent association between situational vulnerability and sleepiness and an inverse association between ethnocultural composition and sleepiness after accounting for important baseline characteristics. Whereas prior studies have explored individual dimensions of vulnerability and their association with OSA, our study is one of the first to take a holistic approach to this analysis, using CIMD as a composite marker of neighbourhood-level social vulnerability. Using this multidimensional index, we were able to account for the intersections of marginalization within our patient populations.

We demonstrated an association between neighbourhood-level situational vulnerability and severity of OSA at the time of referral, which was not robust to adjustment for individual-level BMI. This finding suggests that obesity may mediate the relationship between OSA severity and social vulnerability, which would be supported by a substantial body of literature that has previously linked obesity and OSA, as well as obesity and socioeconomic deprivation^31,32^.

We also observed an inverse relationship between ethnocultural composition and OSA severity, suggesting that individuals residing in regions with ethnically diverse populations may have less severe disease. This finding is in contrast to existing literature which has demonstrated an association between belonging to an ethnic or racial minority and increased severity of OSA^19,33^. A systematic review by Guglielmi et al. found that low SES was a risk factor for the presence of OSA and that disparities in healthcare access, obesity, and SES may mediate the association between racial/ethnic minority status and OSA^16^. Based on the 2016 Canadian Census, the median neighbourhood income for our study population was greater than the provincial median^34^. Moreover, the concentration of ethnic minorities is generally greater within urban centres^35^. Taken together, these findings may suggest that racial or ethnic diversity in the referred study population may be a marker of higher SES which could contribute to this inverse relationship^34^. Furthermore, Canadian populations have higher proportions of immigrant minorities when compared to the United States^36^. Ethnic diversity may be a protective factor in Canadian populations due to the “healthy immigrant” effect which proposes that recent immigrants may be healthier than Canadian-born individuals^36^. However, given the retrospective nature of our study, we cannot exclude the influence of unmeasured confounders or referral bias on this observed association. Further studies assessing the distribution of ethnocultural groups and SES in Alberta would be helpful to better understand this relationship.

We did not observe any significant associations between OSA severity and the dimensions of residential instability and economic dependence. While indicators contributing to residential instability (Table 1) may suggest social vulnerability in some contexts, these may be less accurate measures of social vulnerability in urban areas such as our study setting. Similarly, for economic dependency, our study population lived in areas with higher employment rates and median income compared to the provincial average, resulting in a lower score for this domain (i.e., less disadvantaged). This finding speaks to the potential referral bias within our study population towards more affluent individuals.

Situational vulnerability was significantly associated with sleepiness and severe sleepiness, and residential instability was associated with severe sleepiness. The relationship between social vulnerability and sleepiness has been demonstrated in prior literature^37^, with possible drivers of increased sleepiness in these groups being poorer overall health, higher prevalence of shift work, and higher prevalence of other sleep disorders such as insomnia and circadian rhythm disorders^38^. Sleepiness was inversely related to ethnocultural composition which may relate to differences in SES among FMC Sleep Centre patients from ethnically or racially diverse neighbourhoods compared to other study populations^39^.

This study has several strengths which include its large sample size and robust, validated method of reporting social vulnerability. However, there are also limitations. First, our analysis is only able to capture those patients who were referred to the FMC Sleep Centre for assessment of OSA. Thus, these results do not include individuals who do not access OSA care due to financial, social or medical reasons, and who are often the most vulnerable in society. We also did not capture individuals who were managed independently by their primary care provider or through a community sleep clinic, which may bias our population to more severe/complex presentations. Furthermore, CIMD scores are generated based on 2016 census data and therefore cannot account for evolving community and population dynamics which may impact the ascribed associations.

## Conclusion

In this large, tertiary sleep centre referral population, we found that living in areas with high social vulnerability, measured using a multidimensional index, was associated with OSA severity. These associations, however, appeared to be mediated by BMI. We also found associations between deprivation dimensions and sleepiness that were independent of OSA, which speaks to the broader potential impact of other social and health-related factors on sleepiness. Our findings highlight the complex interplay between social vulnerability and sleep disorders and suggest that composite indices like the CIMD can enhance our understanding of these relationships. The unexpected inverse association between ethnocultural composition and both OSA severity and sleepiness may reflect SES differences between our referral population and patients in other studies. Additional research is needed to explore these relationships further and to assess the generalizability of these findings across different urban settings and populations.

## Data Availability

The data underlying this study cannot be shared due to research ethics board requirements which protect the confidentiality of the research participants.

## References

1. Benjafield, A. V. et al. Estimation of the global prevalence and burden of obstructive sleep apnoea: a literature-based analysis. Lancet Respir. Med. 7, 687–698 (2019).

2. Thompson, C. et al. A portrait of obstructive sleep apnea risk factors in 27,210 middle-aged and older adults in the Canadian Longitudinal Study on Aging. Sci. Rep. 12, 5127 (2022).

3. Yeghiazarians, Y. et al. Obstructive Sleep Apnea and Cardiovascular Disease: A Scientific Statement From the American Heart Association. Circulation 144, (2021).

4. Streatfeild, J., Hillman, D., Adams, R., Mitchell, S. & Pezzullo, L. Cost-effectiveness of continuous positive airway pressure therapy for obstructive sleep apnea: health care system and societal perspectives. Sleep 42, zsz181 (2019).

5. Pendharkar, S. R., Kaambwa, B. & Kapur, V. K. The Cost-Effectiveness of Sleep Apnea Management: A Critical Evaluation of the Impact of Therapy on Health Care Costs. Chest 166, 612– 621 (2024).

6. Povitz, M. et al. Driving consequences of sleepiness in Canadians with obstructive sleep apnea: A population survey. Can. J. Respir. Crit. Care Sleep Med. 6, 298–303 (2022).

7. Pendharkar, S. R. et al. Perspectives on primary care management of obstructive sleep apnea: a qualitative study of patients and health care providers. J. Clin. Sleep Med. 17, 89–98 (2021).

8. Laratta, C. R., Ayas, N. T., Povitz, M. & Pendharkar, S. R. Diagnosis and treatment of obstructive sleep apnea in adults. CMAJ Can. Med. Assoc. J. J. Assoc. Medicale Can. 189, E1481–E1488 (2017).

9. Rizzi, C. F., Ferraz, M. B., Poyares, D. & Tufik, S. Quality-Adjusted Life-Years Gain and Health Status in Patients with OSAS after One Year of Continuous Positive Airway Pressure Use. Sleep 37, 1963–1968 (2014).

10. Johnson, D. A., Ohanele, C., Alcántara, C. & Jackson, C. L. The Need for Social and Environmental Determinants of Health Research to Understand and Intervene on Racial/Ethnic Disparities in Obstructive Sleep Apnea. Clin. Chest Med. 43, 199–216 (2022).

11. Allen, A. J. M. H. et al. Relationship between Travel Time from Home to a Regional Sleep Apnea Clinic in British Columbia, Canada, and the Severity of Obstructive Sleep. Ann. Am. Thorac. Soc. 13, 719–723 (2016).

12. Zhang, H. et al. Factors influencing patient delay in individuals with obstructive sleep apnoea: a study based on an integrated model. Ann. Med. 54, 2816–2828 (2022).

13. Young, T., Evans, L., Finn, L. & Palta, M. Estimation of the Clinically Diagnosed Proportion of Sleep Apnea Syndrome in Middle-aged Men and Women. Sleep 20, 705–706 (1997).

14. Mukherjee, S. et al. An Official American Thoracic Society Statement: The Importance of Healthy Sleep. Recommendations and Future Priorities. Am. J. Respir. Crit. Care Med. 191, 1450–1458 (2015).

15. Etindele Sosso, Fa. & Matos, E. Socioeconomic disparities in obstructive sleep apnea: a systematic review of empirical research. Sleep Breath. 25, 1729–1739 (2021).

16. Guglielmi, O., Lanteri, P. & Garbarino, S. Association between socioeconomic status, belonging to an ethnic minority and obstructive sleep apnea: a systematic review of the literature. Sleep Med. 57, 100–106 (2019).

17. Van Ryswyk, E. M. et al. Primary versus Specialist Care for Obstructive Sleep Apnea: A Systematic Review and Individual-Participant Data-Level Meta-Analysis. Ann. Am. Thorac. Soc. 19, 668–677 (2022).

18. Fatima, D. et al. Access and models of obstructive sleep apnea care: a cross-national comparison of Canadian and Australian patient survey data. J. Clin. Sleep Med. 21, 467–477 (2025).

19. Thornton, J. D. et al. Differences in Symptoms and Severity of Obstructive Sleep Apnea between Black and White Patients. Ann. Am. Thorac. Soc. 19, 272–278 (2022).

20. Johnson, D. A. et al. Influence of neighbourhood-level crowding on sleep-disordered breathing severity: mediation by body size. J. Sleep Res. 24, 559–565 (2015).

21. Spagnuolo, C. M. et al. Distance to Specialist Medical Care and Diagnosis of Obstructive Sleep Apnea in Rural Saskatchewan. Can. Respir. J. 2019, 1–14 (2019).

22. Fatima, D. et al. Exploring patient-borne costs and wait times for obstructive sleep apnea (OSA) care among rural and urban adults. Can. J. Respir. Crit. Care Sleep Med. 7, 21–27 (2023).

23. Franklin, A., Nieri, C., Gallo, N. & Gillespie, M. B. Impact of social vulnerability index on severity of obstructive sleep apnea: Insights from drug-induced sleep endoscopy. Am. J. Otolaryngol. 45, 104450 (2024).

24. Matheson, F. I., Dunn, J. R., Smith, K. L. W., Moineddin, R. & Glazier, R. H. Élaboration de l’indice de marginalisation canadien: un nouvel outil d’étude des inégalités. Can. J. Public Health. 103, S12– S16 (2012).

25. Statistics Canada. The Canadian Index of Multiple Deprivation. (2019).

26. The Canadian Index of Multiple Deprivation: User Guide, 2021. (Statistics Canada = Statistique Canada, Ottawa, 2023).

27. Turan, J. M. et al. Challenges and opportunities in examining and addressing intersectional stigma and health. BMC Med. 17, 7 (2019).

28. Severson, C. A., Pendharkar, S. R., Ronksley, P. E. & Tsai, W. H. Use of Electronic Data and Existing Screening Tools to Identify Clinically Significant Obstructive Sleep Apnea. Can. Respir. J. 22, 215–220 (2015).

29. Tonelli, M. et al. Methods for identifying 30 chronic conditions: application to administrative data. BMC Med. Inform. Decis. Mak. 15, 31 (2016).

30. Postal CodeOM Conversion File Plus (PCCF+) Version 7D, Reference Guide. (2020).

31. Qian, Y. et al. Longitudinal risk factors for obstructive sleep apnea: A systematic review. Sleep Med. Rev. 71, 101838 (2023).

32. Javed, Z. et al. Social determinants of health and obesity: Findings from a national study of US adults. Obesity 30, 491–502 (2022).

33. Stepney, D., Attarian, H. P. & Knutson, K. L. Association between severity of obstructive sleep apnea and its common symptoms varies by race, ethnicity, and sex. J. Clin. Sleep Med. 19, 1727– 1733 (2023).

34. Canadian Income Survery, 2016. (2018).

35. Immigration and Ethnocultural Diversity in Canada - National Household Survey. (2011).

36. Ramraj, C. et al. Equally inequitable? A cross-national comparative study of racial health inequalities in the United States and Canada. Soc. Sci. Med. 1982 161, 19–26 (2016).

37. Groeger, J. A. & Hepsomali, P. Social Deprivation and Ethnicity Are Associated with More Problematic Sleep in Middle-Aged and Older Adults. Clocks Sleep 5, 399–413 (2023).

38. Johnson, D. A., Billings, M. E. & Hale, L. Environmental Determinants of Insufficient Sleep and Sleep Disorders: Implications for Population Health. Curr. Epidemiol. Rep. 5, 61–69 (2018).

39. Gander, P. H., Marshall, N. S., Harris, R. & Reid, P. The Epworth Sleepiness Scale: influence of age, ethnicity, and socioeconomic deprivation. Epworth Sleepiness scores of adults in New Zealand. Sleep 28, 249–253 (2005).

